# Predictors of Arterial Stiffness in Adolescents and Adults with Type 1 Diabetes: A Cross-Sectional Study

**DOI:** 10.1101/2021.07.14.21260516

**Authors:** Kaitlin M. Love, William B. Horton, James T. Patrie, Linda A. Jahn, Lee M. Hartline, Eugene J. Barrett

## Abstract

**Introduction:** Individuals with type 1 diabetes have increased arterial stiffness compared to age-matched healthy controls. Our aim was to determine which hemodynamic and demographic factors predict arterial stiffness in this population.

**Research Design and Methods:** Carotid-femoral pulse wave velocity (cfPWV) was examined in 41 young adults and adolescents with type 1 diabetes without microvascular complications. Two ordinary least squares regression analyses were performed to determine multivariate relationships between cfPWV [log_e_] and: 1) age, duration of diabetes, sex, and HbA1c, and 2) augmentation index (AIx), mean arterial pressure, flow-mediated dilation (FMD), and heart rate. We also examined differences in macrovascular outcome measures between sexes.

**Results:** Age, sex, and FMD provided unique predictive information about cfPWV in these participants with type 1 diabetes. Despite having similar cardiovascular risk factors, males had higher cfPWV compared to females but no differences were observed in other macrovascular outcomes (including FMD and AIx).

**Conclusions:** Only age, sex, and FMD were uniquely associated with arterial stiffness in adolescents and adults with uncomplicated type 1 diabetes. Females had less arterial stiffness and similar nitric-oxide dependent endothelial function compared to males. Larger, prospective investigation is warranted to determine the temporal order of and sex-differences in arterial dysfunction in type 1 diabetes.

**Significance of this study:** *What is already known about this subject?:*

- A measure of central artery stiffness, carotid-femoral pulse wave velocity (cfPWV) predicts renal outcomes, cardiovascular events, and mortality in persons with type 1 diabetes.
- Age, race, mean arterial pressure, waist-to-height ratio/body mass index, presence of microalbuminuria have all been uniquely associated with cfPWV in populations with type 1 diabetes.
- The relationship between cfPWV and flow mediated dilation (FMD), a measure of nitric oxide dependent endothelial function at the brachial artery, has not been defined in type 1 diabetes, and the relationship between diabetes duration and these vascular measures are unclear.

*What are the new findings?:*

- Arterial stiffness and NO-dependent endothelial dysfunction were highly prevalent even within 5 years of diabetes diagnosis (16.7% and 83.3% respectively) in this cohort of adolescents and adults with uncomplicated type 1 diabetes.
- FMD was uniquely associated with cfPWV.
- Augmentation index (AIx), mean arterial pressure, and diabetes duration were not predictive of cfPWV.
- Despite a greater excess cardiovascular risk associated with type 1 diabetes in females compared to males, macrovascular function was no worse in females when examining sex differences. Females had a lower cfPWV and similar AIx and FMD compared to males.

*How might these results change the focus of research or clinical practice?:*

- Larger, prospective clinical investigation is needed in type 1 diabetes to determine the temporal order of and sex-differences in arterial dysfunction.

## Introduction

Large elastic arteries stiffen with accrual of age and other cardiovascular risk factors. This process holds a number of deleterious consequences for the cardiovascular system and major organs, as arterial stiffness is an independent determinant of cardiovascular disease (CVD) risk^1^. Diabetes mellitus (DM) is a cardiovascular risk factor intricately linked to arterial stiffness given that: (1) arterial stiffness is influenced by both hemodynamic forces and extrinsic factors (including hormones, salt, and glucose)^2^; (2) arterial stiffness increases with deteriorating glucose tolerance status^3^ and is associated with insulin resistance^4^; and (3) arterial stiffness is acutely increased during postprandial hyperglycemia in patients with type 2 DM^5^.

People with type 1 DM have significantly increased arterial stiffness (assessed by carotid-femoral pulse wave velocity^6^ [cfPWV] or brachial-ankle pulse wave velocity^7^) compared to age- and sex-matched healthy controls, and this increase occurs independent of traditional cardiovascular risk factors^6 7^. Notably, these changes occur early in the course of disease, as adolescents with type 1 DM exhibit increased arterial stiffness compared to healthy controls^8-11^. Recent prospective cohort studies have also shown that markers of arterial stiffness, including cfPWV and radial artery augmentation index (AIx), predict renal outcomes, cardiovascular events, and mortality in type 1 DM^12 13^. Despite the well-established relationship between arterial stiffness and type 1 DM, few studies have attempted to define the specific factors associated with elevated arterial stiffness in this population^6 14^. To our knowledge, no studies have examined the relationship between brachial artery flow-mediated dilation (FMD; a marker of vascular nitric oxide [NO]-dependent endothelial function) and cfPWV in type 1 DM.

In the current study, we endeavored to clarify the determinants of increased cfPWV in individuals with type 1 DM. Our prespecified hypothesis was that, among demographic and vascular parameters, duration of DM, mean arterial pressure, and AIx would provide unique predictive information about cfPWV in individuals with type 1 DM. We also hypothesized that males and females with type 1 DM would have comparable markers of arterial stiffness (i.e. cfPWV and AIx) and vascular endothelial function (i.e. FMD).

## Research Design and Methods

### Recruitment and Study Population

Following institutional review board approval, we recruited both an adult and an adolescent cohort by public advertisement and direct mailings. An initial telephone interview was conducted with all respondents, and eligible study participants then fasted overnight and presented to the University of Virginia (UVA) Clinical Research Unit (CRU) for a screening visit.

### Clinical Assessment and Initial Screening

All screening visits and study protocols were conducted at the UVA CRU. Each participant gave written informed consent at their initial visit in accordance with the Declaration of Helsinki before screening to verify inclusion/exclusion criteria. Screening included a detailed medical history and physical examination along with fasting measures of complete blood count, comprehensive metabolic panel, lipid panel, hemoglobin A1c, c-peptide, serum pregnancy test, and urine for albumin/creatinine ratio. Adults with type 1 DM met inclusion criteria if they were ≥18 and ≤50 years old, had body mass index <30 kg/m^2^, had hemoglobin A1c (HbA1c) <9.0% (<75 mmol/mol), and had blood pressure <160/90 mmHg at time of screening. Potential participants were excluded from the adult cohort if they were current smokers or had quit smoking <6 months prior, were taking vasoactive medications (e.g. diuretics, statins, etc.) outside of a stable dose of antihypertensive medication, were pregnant (i.e., positive pregnancy test) or nursing, had any known prior microvascular complications due to type 1 DM, had history of cardiovascular, peripheral vascular, or liver disease, had history of ketoacidosis within the previous 12 calendar months, had LDL cholesterol ≥160 mg/dL (4.1 mmol/L), or had serum potassium ≥5.0 mmol/L at time of screening. Adolescents with type 1 DM met inclusion criteria if they were ≥12 and ≤18 years old, had body mass index 18-25 kg/m^2^, and had HbA1c <9.0% at time of screening. Adolescents were excluded if they were current smokers or had quit smoking <6 months prior, were taking vasoactive medications, had LDL cholesterol ≥160 mg/dL, had blood pressure <100/60 mmHg, had pulse oximetry <90%, were pregnant or nursing, had history of cardiovascular, peripheral vascular, or liver disease, or had history of ketoacidosis within the previous 12 calendar months.

### Study Design

We followed the Strengthening the Reporting of Observational Studies in Epidemiology (STROBE) guidelines^15^ to analyze and report this cross-sectional study. All study protocols were approved by the UVA Institutional Review Board. We analyzed baseline measures of cfPWV, AIx, FMD, systolic blood pressure (SBP), diastolic blood pressure (DBP), mean arterial pressure (MAP), and heart rate for each participant. All vascular assessments in this study were measured per expert recommendations^16 17^ by the same trained operator. Study participants were instructed to avoid alcohol, exercise, and caffeine for 24 hours and to fast overnight prior to admission to the CRU.

### Vascular Measures

#### Hemodynamic Function

Clinical hemodynamic assessments were obtained at the initial screening visit. Blood pressure and heart rate were obtained with a GE Dinamap ProCare 400 vital signs monitor (GE Healthcare; Chicago, IL). Mean arterial pressure (MAP) was calculated as: DBP + ((1/3)*(SBP – DBP))^18^.

#### Carotid-Femoral Pulse Wave Velocity (cfPWV)

cfPWV was measured with a SphygmoCor tonometer (AtCor Medical; Naperville, IL) to assess central aortic stiffness. To minimize the effects of sympathetic activity on cfPWV measurements, participants rested supine in a temperature-controlled room for at least 15 minutes prior to measurement. We measured the distance from the suprasternal notch to the carotid pulse and from the suprasternal notch to the ipsilateral femoral pulse. For each cfPWV measure, 10 seconds of carotid and 10 seconds of femoral arterial waveforms were recorded. cfPWV measures were made in duplicate and the mean value was reported. cfPWV intra-observer reliability was also assessed by having the operator record three serial cfPWV measurements on the same subject over a 4-hour period. The coefficient of variation for cfPWV was 3.63%, indicating good intraobserver reliability^19^.

#### Augmentation Index (AIx)

To assess muscular conduit arterial stiffness, we measured AIx noninvasively with a SphygmoCor tonometer. AIx measurements were obtained at the radial artery by the same trained operator with participants lying in the supine position in a temperature-controlled room for at least 15 minutes prior to measurement. AIx was calculated as the difference of the amplitude of the late systolic peak to the early systolic peak divided by the pulse pressure and expressed as a percentage. AIx values were determined for each pulse over a 30-second period, and a mean value was calculated by the device for each patient and corrected for a heart rate of 75 beats per minute.

#### Flow-Mediated Dilation (FMD)

We measured left brachial artery FMD with the EPIQ 7 cardiovascular ultrasound (Philips Medical Systems; Andover, MA) instrument with a linear array probe (L12-3) steadied by a probe-holder as described previously^20^. FMD images were analyzed using Brachial Analyzer (Medical Imaging Applications, LLC; Coralville, IA) edge detection software by study personnel blinded to subject. We assessed FMD intra-observer reliability by having the same trained observer record 8 serial FMD measurements on the same subject over a 4-hour period. The coefficient of variation was 7.41%, indicating good intra-observer reliability^17 19^.

### Biochemical Analyses

Complete blood count, comprehensive metabolic panel, lipid panel, HbA1c, and serum pregnancy tests were assayed at the UVA Clinical Chemistry Laboratory.

### Statistical Analyses

#### Sample Size

The collective study cohort included participants from two separate studies of adolescents and adults with type 1 DM. All persons with type 1 DM from these prior studies were included in the current study.

#### Statistical Methods

A total of two multivariable ordinary least squares (OLS) regression models were constructed to examine the multivariable relationship between cfPWV and potential predictors. The first OLS regression model examined the linear relationships between cfPWV [log_e_] and the multivariable predictor demographic dataset of patient age (years), duration of DM (years), and HbA1C (%). The second OLS regression model examined the linear relationships between cfPWV [log_e_] and the vascular measures multivariable predictor dataset of AIx (%), MAP (mmHg), FMD (% change), and heart rate (beats/min). The OLS regression model type III extra-sum of squares F-tests served as the pivotal quantities for testing the null hypothesis that cfPWV [log_e_] was not uniquely associated with the predictor variable after accounting for the variability in cfPWV [log_e_] that was explained by the remaining set of OLS regression model predictors. A p≤0.05 decision rule served as the null hypothesis rejection rule.

## Results

Demographic and baseline vascular data from the 41 participants are presented in Table 1 and separated by sex. The cohort was 34% female, had mean age of 24 years, mean DM duration of 14 years, and mean HbA1c of 7.9%. Males and females were similar in age, DM duration, HbA1c, blood pressure, BMI, and lipid profile. Males as a group were taller, had lower percent body fat, and higher VO_2max_ compared to females. Regarding vascular measures, AIx and FMD were not significantly different between sexes but cfPWV was higher in males compared to females (indicating greater central aortic stiffness in males).

**Table 1:** Baseline characteristics by sex.

|  | **Combined (n = 41)** | **Females (n = 14)** | **Males (n = 27)** |
| --- | --- | --- | --- |
| Age (years) | 24.2 ± 1.6 | 24 ± 2.8 | 24.3 ± 2 |
| Ethnicity (n [%]) | Non-Hispanic White- 37 (90.2)<br>Black- 4 (9.8) | Non-Hispanic White- 13 (92.9)<br>Black- 1 (7.1) | Non-Hispanic White- 24 (88.9)<br>Black- 3 (11.1) |
| DM duration (years) | 14.5 ± 1.6 | 14.9 ± 2.5 | 14.3 ± 2.1 |
| HbA1c (% / mmol/mol) | 7.9 ± 0.2 / 63 | 7.7 ± 0.4 / 61 | 8.0 ± 0.2 / 64 |
| Urine albumin to creatinine ratio (mg/g) | 14.5 ± 3.2 | 14.9 ± 6.1 | 14.3 ± 3.8 |
| Use of antihypertensive medication (n [%]) | 6 (14.6%) | 1 (7%) | 5 (18.5%) |
| BMI (kg/m<sup>2</sup>) | 24.1 ± 0.6 | 24.6 ± 0.8 | 23.9 ± 0.8 |
| Height (cm) | 173.6 ± 1.5 | 167 ± 1.3 | 177.1 ± 1.9* |
| Weight (kg) | 73.6 ± 2.5 | 68.4 ± 2.9 | 76.3 ± 3.4* |
| Percent body fat (%) | 22.7 ± 1.5 | 29.7 ± 2 | 19 ± 1.6* |
| VO2max (mL/kg/min) | 40.7 ± 1.6 | 32.8 ± 1.3 | 44.9 ± 1.8* |
| Systolic blood pressure (mmHg) | 119.8 ± 2.7 | 112.9 ± 4 | 123.3 ± 3.3 |
| Diastolic blood pressure (mmHg) | 68.8 ± 1.4 | 67.6 ± 2.2 | 69.4 ± 1.8 |
| Total cholesterol (mg/dL / mmol/L) | 160.4 ± 5.4 / 4.15 ± 0.1 | 172.4 ± 8.9 / 4.46 ± 0.2 | 154.2 ± 6.6 / 3.99 ± 0.2 |
| LDL cholesterol (mg/dL / mmol/L) | 98.1 ± 4.8 / 2.5 ± 0.1 | 107.6 ± 7.4 / 2.8 ± 0.2 | 93.2 ± 6.1 / 2.4 ± 0.2 |
| HDL cholesterol (mg/dL / mmol/L) | 51.8 ± 1.3 / 1.3 | 54.4 ± 1.9 / 1.4 | 50.4 ± 1.7 / 1.3 |
| Triglycerides (mg/dL / mmol/L) | 63.1 ± 4.6 / 0.7 ± 0.1 | 62.5 ± 6.6 / 0.8 ± 0.1 | 63.4 ± 6.2 / 0.7 ± 0.1 |
| FMD (% change) | 7.4 ± 0.4 | 8.2 ± 0.8 | 7 ± 0.5 |
| AIx (%) | 3.8 ± 2.1 | 9.1 ± 4.4 | 1.3 ± 2.2 |
| cfPWV (m/sec) | 6 ± 0.3 | 5.3 ± 0.3 | 6.4 ± 0.4** |

Data presented as mean ± SEM. P-values represent Welch's t-tests comparing sexes. \*indicates p < 0.001, \*\*indicates p = 0.03. Abbreviations: DM= diabetes mellitus; HbA1c= hemoglobin A1c; VO2max= maximum oxygen uptake; LDL cholesterol= low density lipoprotein cholesterol; HDL cholesterol= high density lipoprotein cholesterol; FMD= flow-mediated dilation; AIx= augmentation index; cfPWV= carotid-femoral pulse wave velocity.

We examined macrovascular function based on duration of DM. Defining arterial stiffness as cfPWV greater than the 90^th^ percentile for sex and age based on previously defined reference intervals^21^, one of six (16.7%) participants with DM duration <5 years, three of seven (42.8%) participants with DM duration 5 to 10 years, and two of 23 (8.7%) participants with DM duration >10 years met criteria for arterial stiffness. Five participants in the DM duration 5-to-10 years group had cfPWV measures of insufficient quality for analysis. Conversely, there was a higher frequency of NO-dependent endothelial dysfunction (defined as FMD ≤8.1%^22 23^) among our study cohort. Specifically, five of six (83.3%) participants with DM duration <5 years, seven of 12 (58.3%) participants with DM duration 5 to 10 years, and 14 of 23 (60.9%) participants with DM duration >10 years met criteria for impaired FMD. Four participants (2 males/2 females) had elevated urine albumin to creatinine ratio (i.e., >30 mg/g) on screening. All of these participants with microalbuminuria on screening demonstrated impaired endothelial function and one met criteria for arterial stiffness.

Six participants were receiving antihypertension treatment, each with an angiotensin-converting enzyme inhibitor, but this was not significantly different between sexes. Characteristics of these participants are included in the Supplemental Table. Five of six had impaired FMD and one had evidence of central arterial stiffness.

The demographic OLS regression model is presented in Table 2. Sex and age provided unique predictive information about cfPWV, while DM duration and HbA1c did not. All four variables as a collective unit provided significant predictive information about cfPWV.

**Table 2:** Demographic multivariable ordinary least squares regression model for cfPWV.

| Table 2A: OLS regression coefficients for the regression of ln(PWV) onto sex, age, duration of DM and HbA1c |  |  |  |  |
| --- | --- | --- | --- | --- |
| Regression Parameter | Parameter Estimate | Standard Error | Lower 95% CL | Upper 95% CL |
| $\beta_0$ | 0.9896 | 0.3014 | 0.3749 | 1.6043 |
| $\beta_1$ | -0.1435 | 0.0681 | -0.2824 | -0.0046 |
| $\beta_2$ | 0.0190 | 0.0074 | 0.0039 | 0.0341 |
| $\beta_3$ | -0.0051 | 0.0073 | -0.02 | 0.0098 |
| $\beta_4$ | 0.0545 | 0.0299 | -0.0065 | 0.1155 |

| Table 2B: Type III ANOVA F-tests |  |  |  |  |  |
| --- | --- | --- | --- | --- | --- |
| Predictor Variable | Degrees of Freedom | Partial Sum of Squares | Mean Square Error | F-statistic | P-value |
| Sex | 1 | 0.16 | 0.16 | 4.44 | 0.043 |
| Age | 1 | 0.24 | 0.24 | 6.57 | 0.015 |
| Duration DM | 1 | 0.02 | 0.02 | 0.49 | 0.489 |
| HbA1c | 1 | 0.12 | 0.12 | 3.32 | 0.078 |
| TOTAL | 4 | 0.95 | 0.24 | 6.46 | 0.001 |
| ERROR | 31 | 1.14 | 0.04 |  |  |

The vascular OLS regression model is presented in Table 3. In this model, FMD alone offered unique predictive information about cfPWV, while AIx, MAP, and HR did not. All variables as a collective unit did not provide significant predictive information.

**Table 3:** Vascular multivariable ordinary least squares regression model for cfPWV.

| Table 3A: OLS regression coefficients for the regression of ln(cfPWV) onto AIx, MAP, FMD and HR. |  |  |  |  |  |
| --- | --- | --- | --- | --- | --- |
| Regression Parameter | Parameter Estimate | Standard Error | Lower 95% CL | Upper 95% CL |  |
| $\beta_0$ | 1.8677 | 0.4405 | 0.9693 | 2.7661 | |
| $\beta_1$ | 0.0018 | 0.003 | -0.0043 | 0.0079 | |
| $\beta_2$ | 0.0044 | 0.0038 | -0.0034 | 0.0122 | |
| $\beta_3$ | -0.0301 | 0.0144 | -0.0595 | -0.0007 | |
| $\beta_4$ | -0.0034 | 0.0027 | -0.0089 | 0.0021 | |
| Table 3B: Type III ANOVA F-tests. |  |  |  |  |  |
| Predictor Variable | Degrees of Freedom | Partial Sum of Squares | Mean Square Error | F-statistic | P-value |
| AIx | 1 | 0.02 | 0.02 | 0.33 | 0.567 |
| MAP | 1 | 0.07 | 0.07 | 1.34 | 0.256 |
| FMD | 1 | 0.22 | 0.22 | 4.38 | 0.045 |
| HR | 1 | 0.08 | 0.08 | 1.54 | 0.224 |
| TOTAL | 4 | 0.51 | 0.13 | 2.49 | 0.064 |
| ERROR | 31 | 1.59 | 0.05 |  |  |

## Conclusions

Despite similar baseline characteristics and cardiovascular risk factors between sexes, males with type 1 DM had higher cfPWV (indicating greater central arterial stiffness) compared to women with type 1 DM. This was despite male participants having higher VO2max (suggesting greater cardiorespiratory fitness/muscle mass). Prior studies in other populations have shown an association between higher cardiorespiratory fitness and lower cfPWV^24^ and that aerobic exercise training reduces cfPWV^25^. Furthermore, while women typically have lower cfPWV compared to age-matched men, the sex difference observed in our study was more exaggerated than that observed in healthy individuals^21^. More male participants were treated for hypertension in our cohort; however, this was not statically different between sexes and those with hypertension were not disproportionately impacted by elevated cfPWV. Interestingly, there was no sex difference in either FMD or AIx. While larger studies in the type 1 DM population are certainly needed to verify these findings, our initial results are hypothesis-generating. Recent evidence has exposed a twofold excess fatal CVD risk in women with type 1 DM, compared to their male counterparts^26^, though reasons for this are unclear. In the current study, macrovascular function was comparable and aortic stiffness lower in women compared to men. These findings suggest that the increased CVD risk for women with type 1 DM may not be due to differences in macrovascular function. Coronary microvascular dysfunction is increasingly recognized as an important risk factor for CVD events^27^ and is more prevalent in individuals with DM^28^. Haas et al. recently showed that women with type 2 DM and good cardiometabolic control had reduced coronary microvascular function compared to men with type 2 DM^29^. Whether such a sex-based discrepancy in coronary microvascular function exists in type 1 DM, with women potentially having greater dysfunction and this being a primary driver for their increased cardiovascular risk, warrants further investigation.

Contrary to our hypothesis, only age and sex provided unique predictive information about cfPWV in the demographics model. Interestingly, DM duration and HbA1c were not significant predictors. Age and sex are both known to impact cfPWV^21^ and we therefore included these variables in the demographics model to account for their expected influence. One prior study of adolescents with poorly-controlled type 1 DM did find a significant independent association between HbA1c and cfPWV, but as in the current study, DM duration was not significantly associated with cfPWV^30^. The discordant results for HbA1c likely relate to the fact that their study had serial HbA1c measurements available for analysis, while ours included HbA1c measurement at only one time point prior to vascular measurements.

Carotid-femoral PWV is the gold standard estimate of arterial stiffness in adolescents and adults^31 32^ and has a strong independent association with subclinical atherosclerosis^32^. As previously noted, cfPWV is an important risk marker for renal outcomes, cardiovascular events, and mortality in people with type 1 DM^13^, though few prior studies have examined predictors of cfPWV in this population. A multivariable regression model from the larger SEARCH CVD study with a population of 402 adolescents and young adults with type 1 DM and 206 healthy controls found that factors uniquely associated with higher cfPWV included presence of type 1 DM, older age, race other than non-Hispanic white, higher MAP, higher waist-to-height ratio, and presence of microalbuminuria^9^. Another study of 68 individuals with type 1 DM and 68 age- and sex-matched healthy controls found that age, BMI, type 1 DM, and low-grade inflammation predicted cfPWV in men, whereas age, BMI, MAP, and type 1 DM predicted cfPWV in women^6^. Within our vascular model, we found that FMD significantly predicted cfPWV, while HR, MAP, and AIx did not. We were surprised that MAP did not have a significant association with cfPWV in light of the prior results in type 1 DM and the fact that MAP is a known cfPWV determinant^16^. This lack of association likely relates to the fact that the blood pressure used to calculate MAP was obtained at the initial screening visit (up to 4 weeks prior to vascular studies). We used MAP obtained at this time, instead of study admission, to assess the reliability of MAP measured in a clinical environment (e.g., an ambulatory office setting) to predict arterial stiffness, and a change in blood pressures/MAP across this time is possible.

We also did not observe a significant association between AIx and cfPWV. In type 1 DM, both AIx and cfPWV are elevated^33^ and predict cardiovascular events and mortality^12 13^. The lack of association we observed may relate to intrinsic differences between AIx and cfPWV as measures of vascular stiffness. Whereas AIx is a peripheral arterial measurement influenced by numerous determinants including heart rate and contractility, cfPWV is a more direct measurement of central aortic and aorto-iliac artery stiffness, and cfPWV is considered the “gold-standard” non-invasive measure of arterial stiffness ^34^. In young healthy individuals, central arteries show greater elasticity compared to more muscular peripheral arteries, and central arteries are preferentially stiffened by aging and hypertension^35^. In type 1 DM, a dissociation between central and peripheral arterial stiffness, as evaluated by cfPWV and AIx, was also previously reported with a lack of association between the two measures after adjusting for cardiovascular risk factors^36^. Taking into account our findings, this may suggest a more important role for cfPWV measurement in early cardiovascular risk stratification for type 1 DM.

That only FMD provided unique predictive information about cfPWV was surprising given that FMD measures brachial artery NO-dependent endothelial function. The endothelium releases the potent vasodilator NO in response to shear stress induced by reactive hyperemia elicited by FMD^37^ testing. NO bioavailability influences dynamic changes in the arterial wall^38^, and preserved endothelial NO production is an important atheroprotective factor. FMD specifically has a strong inverse linear relationship with cardiovascular events and mortality. In fact, every 1% increase in FMD correlates with a 9% reduction in cardiovascular events^39^. Impaired vasodilation measured by FMD is commonly seen early in the disease process of persons with type 1 DM, as others have reported ∼36% of children and adolescents already have reduced FMD within 5 years of diagnosis^22 40^. Despite reasonable glycemic control at the time of study, our population had an even greater prevalence of NO-dependent endothelial dysfunction that was relatively stable across groups with longer DM duration. The consistent finding of early impaired NO-dependent endothelial function and the association between FMD and HbA1c has led some to hypothesize that there is a metabolic memory phenomenon in type 1 DM whereby early glycemic control is highly formative of endothelial function^22^, although more data are needed to validate this hypothesis. This may explain why we saw no significant relationship between HbA1c at time of study and cfPWV.

To our knowledge, the current study is the first to investigate the specific association between cfPWV and FMD within type 1 DM. A prior study of 68 individuals with type 1 DM and 68 sex- and age-matched healthy controls reported that endothelial dysfunction was more frequent in the type 1 DM cohort but not associated with cfPWV after adjusting for potential confounders^41^. However, the study used reactive hyperemia peripheral arterial tonometry (RH-PAT) to assess endothelial function and it is noteworthy that RH-PAT results failed to significantly correlate with FMD in either the Framingham Heart Study’s Offspring, Third Generation, or Omni cohorts^42^. Moreover, brachial artery FMD is recognized as the gold standard for non-invasive assessment of endothelial function^43^. While FMD was a significant predictor of cfPWV in our type 1 DM population, associations between cfPWV and FMD in other populations are inconsistent. In newly-diagnosed hypertensive individuals (n=189), FMD had a significant independent association with carotid intimal thickness but not cfPWV^44^. Similarly, in the Young Finns study (n= 1754), FMD did not modulate the association between cardiovascular risk factors and cfPWV in young adults^45^. Among individuals with type 2 DM, a significant independent association was found between brachial-ankle cfPWV and FMD but only in individuals with less advanced atherosclerosis (i.e., lesser carotid intima-media thickness)^46^. Others found that FMD was inversely associated with, but not a unique predictor of, cfPWV in individuals with long-standing type 2 DM and hypertension^47^. Still, the association between cfPWV and FMD in our type 1 DM population raises the possibility that dysglycemia or perhaps insulin resistance, even in the absence of other cardiovascular risk factors, increases the risk of both vascular endothelial and arterial wall dysfunction. Along these lines, a recent meta-analysis of 58 studies examining endothelial function in persons with type 1 DM (n= 2,322) and healthy controls (n= 1,777) corroborated the presence of early endothelial and vascular smooth muscle dysfunction in children and adults with type 1 DM^48^. The study also reported that endothelial dysfunction seems to be more pronounced within macrovascular than microvascular beds, fostering the debate on their relative temporal appearance.

While our study provides a unique look at macrovascular function in adolescents and adults with uncomplicated type 1 DM, it contains several important limitations. First, it is a small cross-sectional examination across a wide age spectrum. Second, it defines neither the time sequence nor the ontology of macrovascular dysfunction. Third, this study includes HbA1c at only one time point. Serial HbA1c measures would provide valuable data when evaluating long-term vascular function. With these limitations in mind, the similar representation of cardiovascular risk factors between males and females in this cohort provides valuable information about early macrovascular function across sexes in type 1 DM prior to the advent of microvascular or macrovascular complications.

In conclusion, age, sex, and FMD were the only factors uniquely associated with cfPWV in our cohort of adolescents and adults with uncomplicated type 1 DM. Males had higher cfPWV, indicating greater central arterial stiffness, but similar FMD and AIx to females. Endothelial dysfunction was highly prevalent among our participants regardless of DM duration. Further study is warranted to determine the generalizability of our results and define the sequence of and sex-differences in macrovascular dysfunction present in type 1 DM.

## Supporting information

STROBE checklist

## Data Availability

Data contained in this manuscript is available upon request.

## Contributions Statement

K.M.L. and W.B.H: study planning, data analysis, manuscript preparation; J.T.P.: data analysis methods, data analysis, manuscript preparation; L.A.J: subject recruitment, study execution, data analysis and manuscript preparation; L.M.H.: subject recruitment, study execution, manuscript preparation; E.J.B.: study planning, execution, data analysis, manuscript preparation and funding acquisition.

## Funding

This work was supported by research grants from the NIH (DK101944 and DK073059) to EJB and in part by the National Center For Advancing Translational Sciences of the National Institutes of Health under Award Numbers KL2TR003016/ULTR003015 (to KML and WBH as iTHRIV scholars). The content is solely the responsibility of the authors and does not necessarily represent the official views of the National Institutes of Health.

## Competing interests

None to report.

## Ethics approval

The studies included in this manuscript were reviewed and approved by the institutional review board of the UVA Health System.

## Notes

### Competing Interest Statement

The authors have declared no competing interest.

### Author Declarations

All study protocols included in this cross-sectional study were approved by the University of Virginia Institutional Review Board for Health Sciences Research.

