## Supplemental Table for "Predictors of Arterial Stiffness in Adolescents and Adults with Type 1 Diabetes: A Cross-Sectional Study"

**Supplemental Table: Descriptive statistics of individuals treated for hypertension**

| Characteristic | Mean ± SEM | 95% Confidence Interval |
|---|---|---|
| Age (years) | 36 ± 3.2 | 28 – 45 |
| Diabetes duration (years) | 27 ± 3.5 | 18 – 36 |
| Presence of elevated urine albumin to creatinine ratio (n) | 1 | |
| Systolic blood pressure (mmHg) | 143 ± 6 | 127 – 158 |
| Diastolic blood pressure (mmHg) | 78 ± 2.2 | 72 – 83 |
| Flow mediated dilation (%) | 7 ± 0.9 | 4.7 – 9.3 |
| cfPWV (m/sec) | 7.5 ± 1 | 4.9 –10 |
| cfPWV percentile | 53 ± 15 | 13 – 93 |
| AIx (%) | 14 ± 6.5 | -2 – 31 |

Abbreviations: ACE = Angiotensin-converting enzyme; AIx= augmentation index; cfPWV= carotid-femoral pulse wave velocity
